# Efficient Prevalence Estimation and Population Screening for SARS-CoV-2

**DOI:** 10.1101/2020.05.18.20105882

**Authors:** Kenneth Gundersen, Jan Terje Kvaløy, Håkon K. Gjessing, Iren Høyland Löhr

**Author notes:** Corresponding author: Jan Terje Kvaløy PhD, Department of Mathematics and Physics, University of Stavanger, 4036 Stavanger, Norway.

## Abstract

The testing capacity for SARS-CoV-2 using RT-PCR analyses can be substantially increased by using pooling techniques. Here we discuss two important consequences of this. First, prevalence estimation can be done with a very limited number of RT-PCR analysis. Second, for screening purposes, the number of detected cases can largely increased.

## Introduction

The most common testing method for SARS-CoV-2 is RT-PCR analysis of nasopharyngeal and/or throat swab samples. In most countries, testing capacity is highly limited due to lack of equipment and necessary testing reagents. Consequently, some laboratories have explored pooling techniques to increase the number of individuals tested with the available number of tests.^1^ At Stavanger University Hospital we have operationally verified the possibility of pooling eight samples with maintained diagnostic sensitivity. We would like to draw the attention to two, maybe less obvious, opportunities enabled by pooling of up to 32 or even more samples that appear to be possible within the PCR protocol recommended by WHO for SARS-CoV-2 detection.

## SARS-CoV-2 prevalence measurement with minimal number of PCR tests

Determination of the prevalence of SARS-CoV-2 in the population is of high importance for a number of reasons. For instance, the “true” prevalence is a crucial input to epidemiologic modelling, and for monitoring the effect of social distancing and other measures to suppress spread of SARS-CoV-2.^2^ By single-step pooled analysis, the number of PCR analyses necessary is reduced by a factor equal to the number of samples in each pool. An estimate of the prevalence can be calculated from the proportion of positive pools; it is not necessary to identify the exact number of positive samples in each pool. The technique is well described in the literature^3-5^ and corrects for the possibility that there is more than one positive sample in each positive pool. Details about tools and methods are given in the supplementary material. To further reduce the number of required PCR analyses, a higher degree of pooling can be employed, even if the corresponding reduction in sensitivity would be considered unacceptable for diagnostic purposes. For prevalence estimation the reduced sensitivity is, however, straightforward to correct for, as long as it is known. Yelin et al.^1^ have indicated that with pooling of 32 samples the sensitivity is 90% (10% false negative rate), relative to that of single sample analysis. With this degree of pooling it would take 200 PCR tests to estimate the prevalence from a dataset of 6400 individual samples. Figure S1 in the supplementary material indicates the 90% confidence intervals for various sample sizes, degree of pooling and prevalences.

## Maximizing the number of detected cases when screening in a low prevalence population

The most common purpose of pooled testing is to analyze as many samples as possible with a given laboratory capacity and maintain a diagnostic level of sensitivity. Further, to combat the current covid-19 epidemic many countries are adapting a “suppression” strategy as recommended by Ferguson et al. ^2^ and WHO.^6^ The basic idea of this strategy is, after a period of aggressive social distancing measures, to do extensive testing, case tracing and isolation to keep the prevalence at a low level until a vaccine becomes available. In this context it is important to maximize the number of infected people detected with the available testing resources, rather than to insist that all tests from the community setting are being performed with the highest level of sensitivity. Currently, the limited literature indicates that with a pool size of 32 the sensitivity drops to around 90%.^1^ When used for population screening pooling of 32 samples will nevertheless allow for detection of far more infected people than for the same number of PCR tests if using one test per person. If screening a population with a prevalence of 0.005 and using pooling of eight samples a mean of around 6000 people can be screened and a mean of 30 cases detected using 1000 PCR tests. By pooling 32 samples, however, around 12000 people can be screened and a mean of 55 cases detected with the same 1000 PCR tests. Figure S2 shows the corresponding numbers for a range of prevalence values in the screened population. It appears that if using this approach some high-income countries already would have the PCR capacity needed to perform extensive screening of the public, if the prevalence is first suppressed to a low level with strong social distancing measures.

Limitations of both of the above suggested applications of extended pooling is that while the required number of PCR analysis is reduced, the number of samples to be collected is unchanged. Further, the relationship between pool size and test sensitivity is not yet fully established and might also vary between individual laboratories and various PCR protocols, and therefore have to be verified locally.

## Transparency declaration

The authors have no conflicts to declare. No external funding relevant for this work was received by any of the authors.

## Data Availability

No data are reported in the article

## Supplementary Material

This supplementary file first provides the plots referenced in the main text. The calculations behind the plots are explained at the end of the document. The R code used to generate the plots can be found in the GitHub repository: https://github.com/jtkgithub/SARSCoV2_pooled_testing.

### Prevalence estimation

Figure S1 shows 90% confidence intervals for the estimated prevalence obtained for different sample sizes (number of persons sampled), degrees of pooling, and prevalences. Figure S3 below shows the sensitivity assumed in these calculations, depending on the degree of pooling.

**Figure S1:**
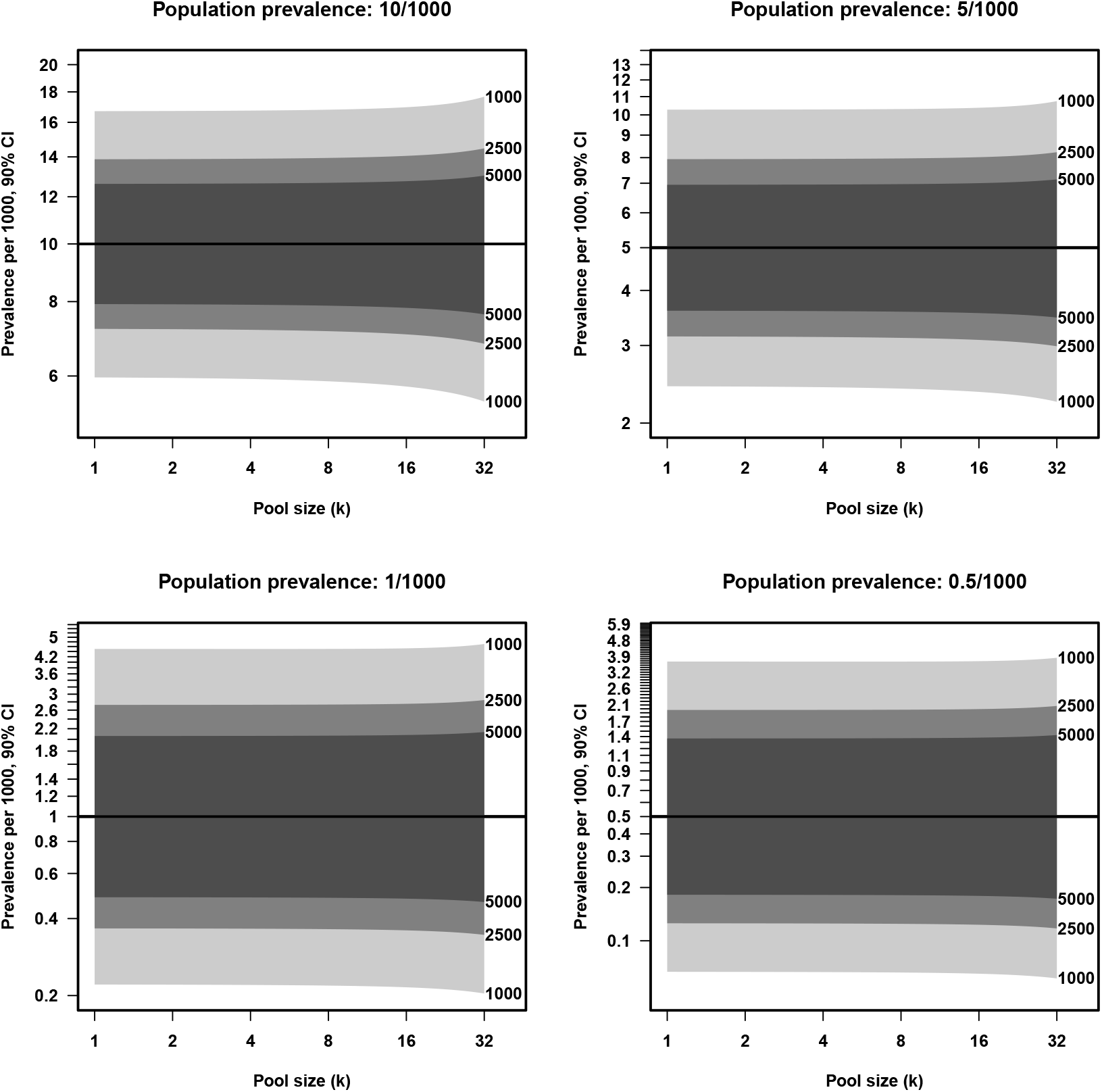
Precision (90% confidence intervals) of prevalence estimates based on pooled sampling, depending on prevalence, degree of pooling, and the total number of persons sampled (shown in right hand column of the plot). Note that the number of pools needed to be analyzed is the number of persons divided by the degree of pooling. The calculations assume a pool analysis sensitivity as shown in Figure S3. See text for further description.

### Screening

Figure S2 shows how many persons can be included in the screening and the expected number of infected persons identified for three different screening strategies. The first strategy is to do no pooling. The second strategy is to first test samples in pools of 8. For the pools that are positive, samples from all 8 persons are tested individually. In the third strategy samples are first tested in pools of 32. For pools that are positive a second test in 8 pools of size 4 are made. Then finally for the positive pools of 4, individual tests are run.

**Figure S2:**
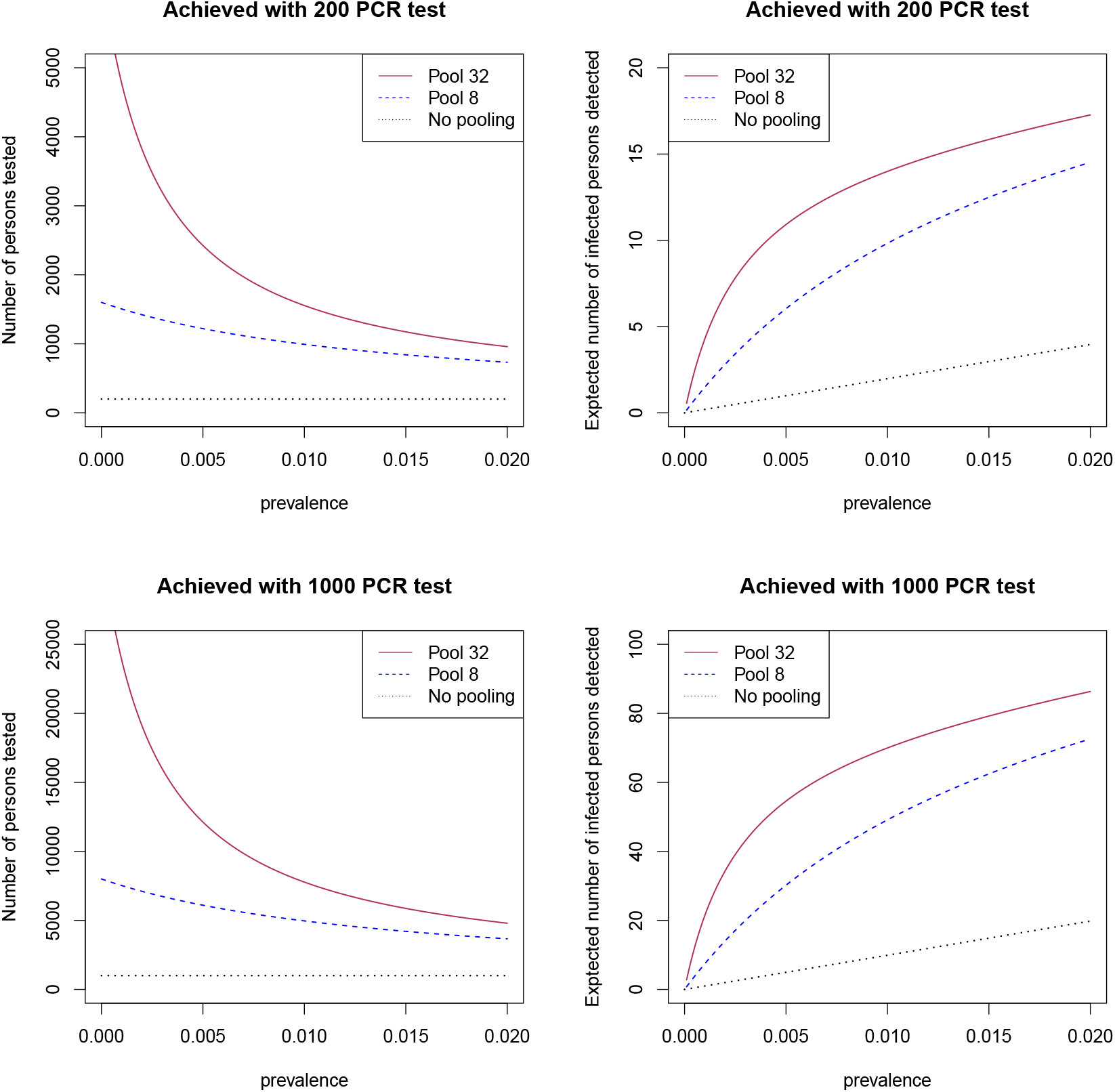
The left plots depicts the number of persons that could be tested and the right plots the expected number of infected persons that would be identified with two different PCR budgets and three different test strategies: 1) No pooling; 2) Pool size 8 in the first testing and re-testing of all samples in the positive pools; 3) Pool size 32 in the first testing, re-testing in pools of 4 for the positive pools, re-testing all for the positive pools of size 4.

In the illustrations shown in Figure S2 we have assumed a sensitivity of 99% for pools of size 8 and a sensitivity of 90% for pools of size 32. Similar plots for other scenarios can be generated using the R code in the GitHub repository. Note that both the numbers of persons included and the expected number of cases identified scale linearly with the number of available PCR analyses. For 1000 analyses the numbers are five times that of 200 analyses.

**Figure S3:**
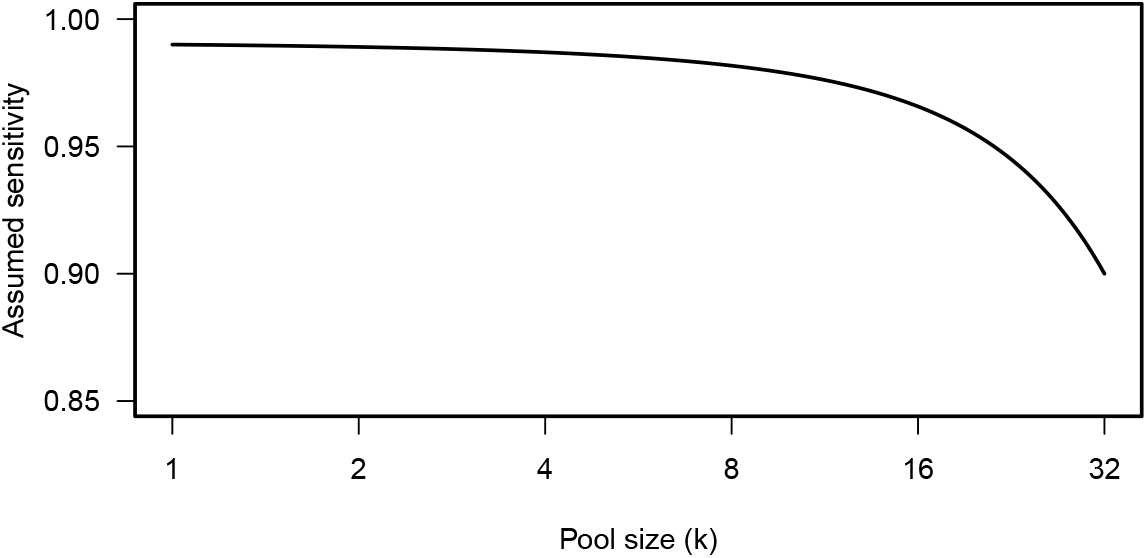
Sensitivity of pooled samples, as assumed in the confidence interval calculations for Figure S1

### Mathematical details

Let p be the true prevalence, *k* the number of samples merged in one pool and *π*(*k*) the probability that a pool of *k* tests is positive. Let further *s*(*k*) be the sensitivity of the test for a pool of *k* tests. We assume the specificity of the test to be 1. Then

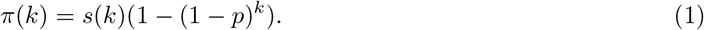

#### Prevalence estimation

For prevalence estimation, *p*, we first estimate *π*(*k*) by

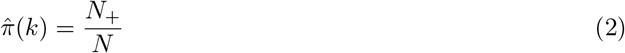

where *N*_+_ is the number of positive pools and *N* is the number of pools. The total number of persons tested is *n_pers_* = *kN*. From (1) and (2) we get that

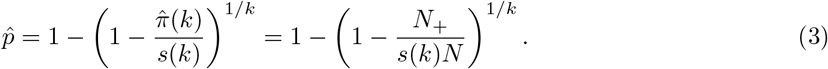

Moreover, to obtain a confidence interval for 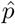 we propose to first calculate Wilson confidence intervals^1^ for 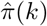 and map this to confidence limits for 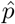 by inserting the upper and lower limits in (3). Wilson confidence intervals are available from the `binom.confint` function in the `R package binom`^2,3^. Additional statistical tools for computing prevalences from pooled samples can be found in the `prevalence` package ^4,5^.

#### Screening strategies

For considerations around the screening strategies let first *N_PCR_* be the total number of PCR analyses which needs to be performed and let *N_PCR_*_,1_ be the number of PCR analyses run in the first round. With no pooling *N_PCR_* = *N_PCR_*_,1_. For pooling stratgies *N_PCR_* will be a random variable since we do not know how many positive pools we will find. For the strategy with pooling of 8 samples we have

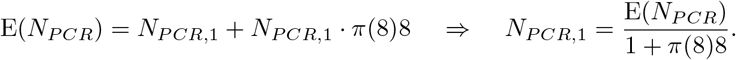

By inserting the budget of available PCR analyses for E(*N_PCR_*) (e.g. 200 and 1000 in Figure S2) we get that the total number of persons that can be tested is *n_pers_* = 8*N_PCR_*_,1_. Let *n_pos_* be the total number of identified positive cases, and let *n_pos_*_,8_ denote the number of positive cases in a pool of 8. Then

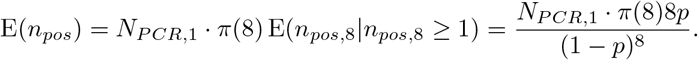

For the strategy starting with 32 pools then re-testing in pools of 4 if positive and finally testing all samples in positive pools of 4, similar calculations give.

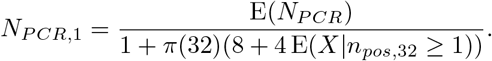

Here *X* is the number of positive pools among eight pools of size 4, and 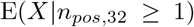 can be approximated by

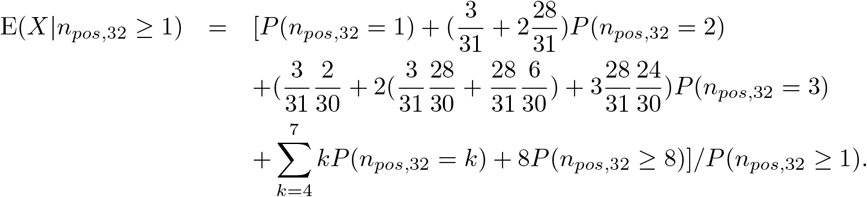

The last two terms 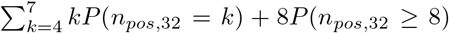 implies that this is an upper bound, the approximation being very accurate for small *p*. Finally, we get the expected number of detections by

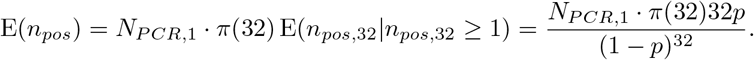

## Acknowledgements

We thank Eva Bernhoff and Ola Brønstad Brynildsrud for useful discussions and contributions.

## References

1 Yelin I, Aharony N, Shaer-Tamar E, et al. Evaluation of COVID-19 RT-qPCR test in multi-sample pools. *medRxiv* 2020; published online March 27. DOI:10.1101/2020.03.26.20039438 (preprint).

2 Ferguson NM, Laydon D, Nedjati-Gilani G, et al. Report 9: Impact of non-pharmaceutical interventions (NPIs) to reduce COVID19 mortality and healthcare demand. Imperial College COVID-19 Response Team, 2020, https://www.imperial.ac.uk/media/imperial-college/medicine/mrc-gida/2020-03-16-COVID19-Report-9.pdf (accessed March, 20).

3 Speybroeck N, Williams CJ, Lafia KB, Devleesschauwer B, Berkvens D. Estimating the prevalence of infections in vector populations using pools of samples. Med Vet Entomol 2012; 26: 361-71.

4 Liu A, Liu C, Zhang Z, Albert PS. Optimality of group testing in the presence of misclassification. Biometrika 2012; 99: 245-51.

5 Warasi MS, Tebbs JM, McMahan CS, Bilder CR. Estimating the prevalence of multiple diseases from two-stage hierarchical pooling. Statist Med 2016; 35: 3851-64.

6 WHO. Global surveillance for COVID-19 caused by human infection with COVID-19 virus: Interim guidance 20 March 2020. World Health Organization, 2020. https://apps.who.int/iris/handle/10665/331506 (accessed Apr 6, 2020).

## References

1 Agresti A, Coull BA. Approximate is Better than “Exact” for Interval Estimation of Binomial Proportions. The American Statistician. 1998;52(2):119–126.

2 R Core Team. R: A Language and Environment for Statistical Computing. Vienna, Austria; 2019.

3 Dorai-Raj S. binom: Binomial Confidence Intervals For Several Parameterizations; 2014. R package version 1.1–1.

4 Devleesschauwer B, Torgerson P, Charlier J, Levecke B, Praet N, Roelandt S, et al. prevalence: Tools for prevalence assessment studies.; 2015. R package version 0.4.0.

5 Speybroeck N, Williams CJ, Lafia KB, Devleesschauwer B, Berkvens D. Estimating the prevalence of infections in vector populations using pools of samples. Medical and Veterinary Entomology. 2012 Dec;26(4)361–371

